# The association between adverse experiences throughout the life-course and risk of dementia in the English Longitudinal Study of Ageing

**DOI:** 10.1101/2025.06.20.25329995

**Authors:** Katherine Taylor, Laura D Howe, Rebecca E Lacey, David Carslake, Emma Anderson, Naaheed Mukadam

## Abstract

**Introduction:** Studies investigating the association between adverse experiences across the life-course and dementia consider a narrow range of experiences and use sum scores, assuming each experience has the same impact on dementia risk. We considered the timing, type and cumulation of adverse experiences.

**Methods:** The English Longitudinal Study of Ageing measured adverse experiences in a retrospective interview. Cox proportional hazard models were used to investigate associations between dementia and sum adversity scores, individual experiences, and broad categories adapted from existing frameworks.

**Results:** Number of adult, but not total or childhood, adverse experiences was associated with dementia. Child abuse and adult economic hardship were associated with a 74% and 32% higher hazard of dementia respectively.

**Discussion:** Adulthood adverse experiences associate with dementia in a cumulative risk manner. In childhood, only abuse was associated with dementia. Use of sum scores to summarise adverse experiences throughout the life-course may oversimplify associations with dementia.

## 1. Background

Dementia is a growing public health problem, predicted to cost the UK £90 billion by 2040 [1]. It is estimated that up to 45% of dementia cases are preventable, and research into potentially modifiable risk factors is vital [2].

Adverse experiences throughout the life-course have been hypothesised to increase risk of dementia, impacting individuals on a biological and behavioural level [3–5].

Stress in early-life is thought to be more impactful than stress in mid-life and most literature focusses on the association between dementia and adverse childhood experiences (ACEs)[6]. Meta-analysis revealed that exposure to two or more ACEs, compared to one or fewer, was associated with 1·35 times higher odds of dementia (95% CI:1·20-1·52)[7]. However, all included studies used cumulative risk approaches which assume all ACEs contribute equally to increasing dementia risk [8]. Analysis investigating the effect of individual ACE measures on dementia risk suggests this assumption is likely to be violated [5,7,9,10]. That said, considering ACEs separately does not consider the correlation between them; many experiences are not independent of one another.

Few studies consider the impact of adverse experiences across the whole life-course on dementia risk. Studies have observed mid-life stress to be associated with dementia, with a greater number of stressors and persistent stress exposure being associated with a higher prevalence and incidence of dementia [6,11,12]. A systematic review found that total number of adverse events throughout the life-course were associated with all-cause dementia, reporting a hazard ratio (HR) of 1·21 (95% CI:1·03-1·42). However, within this meta-analysis, only seven studies were included, and many were limited in scope considering only one type of adverse experience [3]. Additionally, multiple studies have been conducted using the UK Biobank which is not representative of the general UK population and contains relatively few dementia cases due to the age of the sample [13,14].

In summary, existing literature is limited in scope, considering solely cumulative measures of adversity or focussing on a narrow range of adverse experiences, using unrepresentative samples, often with short follow-up periods and limited dementia cases. Additionally, most studies focus on ACEs not adversities throughout the whole life-course. This study aims to these address gaps by estimating the relationships between a large collection of adverse experiences and dementia incidence in a sample which is more representative of the UK population. Through consideration of the timing, type and cumulation of adverse experiences we can better elucidate which experiences drive associations between adverse experiences and dementia and gain a greater understanding of their risk mechanism.

## 2. Methods

### 2.1 Study population

Data were obtained from the English Longitudinal Study of Ageing (ELSA; www.elsa-project.ac.uk), a large-scale longitudinal panel study of English adults aged ≥50. Full details of the study population are included in the online supplement and have been published elsewhere [15]. Briefly, participants were interviewed every two years and had a health examination every four years; we used data up to the interview in October 2022.

### 2.2 Sample selection

Following data collection at wave three (2007, mean age 65·3 years), participants undertook a life history interview which asked about previous adverse events.

Participants eligible for inclusion had completed the life history interview and the adverse life events questionnaire (Figure 1). Analyses were restricted to ‘core members’, defined as age-eligible members interviewed at wave one or wave three. Those with dementia at wave three, or who retrospectively reported dementia diagnosis before their life history interview, and those with missing dementia data post wave three were excluded.

**Figure 1.**
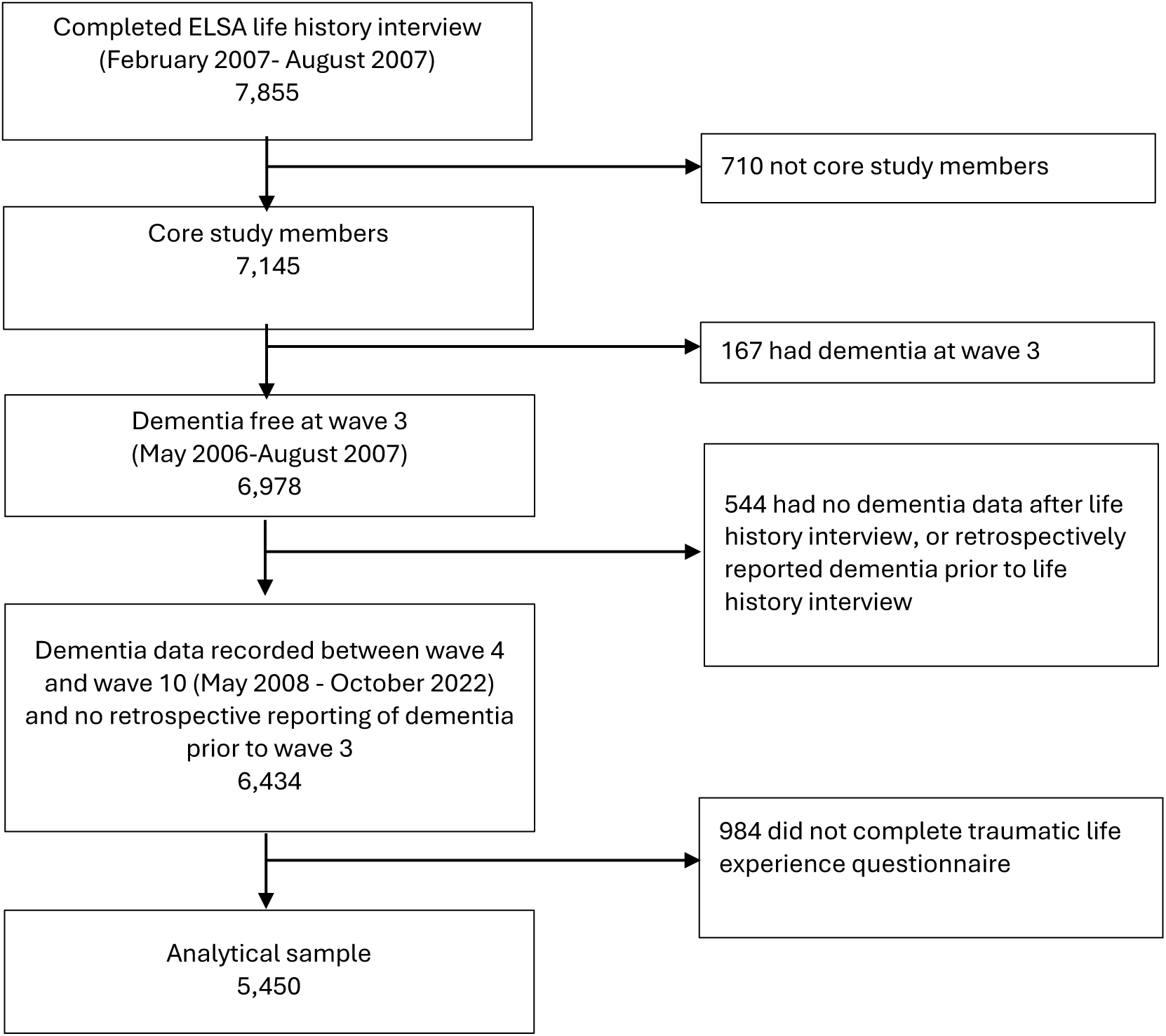
STROBE diagram

### 2.3 Variables

#### 2.3.1 Exposure

Adverse life experiences were assessed at the life history interview. Table 1 outlines the adversity variables considered in our analyses; both individual adversity measures and broad adversity categories derived from those individual measures. Full details on how adverse life experience variables were derived from participants’ answers to questions, and how we decided which adverse experiences to group into broad categories, are provided in the online supplement (Appendix 1). Childhood adverse experiences were defined as those occurring prior to age 16, and adult adverse experiences after age 16. Where possible, the same experiences were considered in adulthood and childhood to investigate the importance of timing of adverse events. Additional adult adversities which are unlikely to occur as a child (such as having had a partner/husband/wife/child addicted to drugs or alcohol) were also incorporated. Sum scores counting the total number of adverse experiences, total number of childhood adversities and total number of adulthood adversities were derived [16,17].

**Table 1.** Broad adverse experience categorisations.

| **Adversity in childhood** | **Adversity in adulthood** |
| --- | --- |
| **Exposure to war/collective violence [17]**<br>• Lost a very close friend or relative in war or military combat aged under 16 | **Exposure to war/collective violence [17]**<br>• Lost a very close friend or relative in war or military combat aged 16 or over<br>• Ever fired a weapon in combat or be fired upon<br>• Witnessed serious injury/death of someone in war or military combat<br>• Have lived in a prisoner of war camp |
| **Economic hardship**<br>• While under the age of 16 parents unemployed for more than 6 months when they wanted to work<br>• Experienced severe financial hardship aged under 16 | **Economic hardship**<br>• Experienced severe financial hardship aged 16 or over<br>• Have been homeless for one month or more |
| **Illness, accidents and natural disasters**<br>• Lost a very close friend or relative due to illness or injury aged under 16<br>• Witnessed an accident/violence which caused death or serious injury (not war) aged under 16<br>• Experienced a natural disaster aged under 16<br>• Had a life-threatening illness or accident aged under 16 | **Illness, accidents and natural disasters**<br>• Witnessed an accident/violence which caused death or serious injury (not war) aged 16 or over<br>• Had a life-threatening illness or accident aged 16 or over<br>• Experienced a natural disaster aged 16 or over |
| **Household challenges [18]**<br>• While under the age of 16 parents abused substances or had a mental illness<br>• Spent most of childhood in a social care setting | **Household challenges [18]**<br>• Have a child who has died<br>• Had a partner/husband/wife/child addicted to drugs or alcohol |
| **Abuse [18]**<br>• Sexual assault aged under 16<br>• Physically abusive parents aged 16 or under | **Abuse [18]**<br>• Sexual assault aged 16 or over<br>• Victim of serious physical attack aged 16 or over |
| • Victim of serious physical attack aged under 16 |  |

#### 2.3.2 Outcome

All-cause dementia, the main outcome measure, was algorithmically defined as previously described [19,20]. Dementia was defined as having self-reported doctor-diagnosed dementia (N=270), scoring 3·6 or higher on the Informant Questionnaire on Cognitive Decline (N=28)[21] or having declined in two or more cognitive domains and a non-transient impairment in one or more activity of daily living (N=217)[20].

Full details of the dementia variable, including how date of diagnosis was derived, are in the online supplement (Appendix 2).

#### 2.3.3 Covariates

Age and sex were self-reported at the life history interview. Ethnicity was reported at wave three and binarized into White or Other ethnicities due to low sample numbers for the latter. Number of books in the home aged ten was used as a proxy measure of childhood socioeconomic status (SES) and binarized into ten books or fewer, and more than ten books. Childhood economic hardship, as defined in Table 1, was considered as both an effect modifier and an adverse experience. We considered it to be distinct from childhood SES, as it focuses on the hardship associated with a low income rather than SES itself.

### 2.4 Statistical Analysis

#### 2.4.1 Primary Analysis

Analysis was performed in RStudio (Version 2023.09.1+494). We used multiple imputation through chained equations to impute missing data. Further details can be found in the supplement (Appendix 3). Multiply imputed data was used for our primary analysis. Sensitivity analyses were conducted using complete case data.

Tetrachoric correlation matrixes among individual adversities and broad adverse experience categories were estimated. We used Cox proportional hazards models to estimate associations between dementia incidence and sum scores of adversities.

Subsequently, Cox models were used to estimate the association between dementia incidence and each exposure separately. This analysis was repeated for broad adversity categories. The study start date was set to the life history interview date.

The time from study start to dementia diagnosis or censoring, defined as a participants last interview where data on dementia was collected, was taken for each participant. Follow-up was left truncated at age at life history interview and age used as the time axis in survival models. All Cox models were adjusted for ethnicity, sex and childhood SES. The proportional hazards assumption was assessed in each model by testing for the independence between residuals and time.

Three Cox models were used to investigate heterogeneity in the hazard of dementia associated with each adversity measure. In addition to the covariates previously included, the models adjusted for 1) the five broad childhood adversity categories defined in Table 1, 2) the five broad adulthood adversity categories defined in Table 1, and 3) all childhood and adulthood broad adversity categories defined in Table 1 combined. Linear hypothesis testing was used to test for heterogeneity among the HR associated with the different adverse experience categories within the same model. Broad categories of adverse experience were used as opposed to individual measures to increase the statistical power of this analysis, as many adverse experiences were uncommon.

Sex and childhood economic hardship were considered as potential effect modifiers [10,22]. Cox models were re-run introducing an interaction term between sex and sum adversity scores, then an interaction term between sex and broad categorisations of adverse experience. This was repeated introducing interaction terms between the same adversity measures and childhood economic hardship. For these analyses, sum adversity scores were adjusted to remove the individual measures used to derive childhood economic hardship.

#### 2.4.2 Sensitivity analysis

Informative censoring may impact the results of the Cox models generated because individuals with dementia may be more likely to drop out of the study before dementia can be reported. Additionally, those with a higher number of adverse experiences have shorter follow-up times and are more likely to be censored before wave ten. We conducted a series of regression models to investigate the potential for informative censoring. Full details of these analyses are in the supplement (Appendix 4) [23]. Analyses using sum scores of adversities were repeated categorising adversity scores into 0,1,2,3,4+ to test for threshold effects. Analyses looking at the association between sum adversity scores and broad adversity categories and dementia were re-run on complete case data.

## 3. Results

Our analytical sample consisted of 5,450 individuals (Figure 1). Excluded individuals were older, more likely to be female, had lower levels of education and were less likely to be White (Supplementary Table 2).

44% of the analytical sample was male and 56% female. Most participants were White (98·4%). The mean age at the life history interview was 66.1 years and average follow-up time was 10.1 years. 515 (9·4%) people developed dementia during the follow-up period. There were positive correlations among the individual adversities which feed into the same broad adversity category and negative correlations between the equivalent adverse experiences in adulthood and childhood. This is due to how experiences were recorded in ELSA; if individuals reported an experience in childhood, they couldn’t report it in adulthood. (Supplementary Figure 1). There were correlations among broad categories of adversity. For example, childhood household challenges were correlated with childhood abuse (r:0·4) (Supplementary Figure 2).

Participants who developed dementia were older (74·0 compared to 65·3) and had a higher number of total adverse experiences (1·6 compared to 1·4). This was driven by a higher number of adult adversities among those with dementia (1·0 compared to 0·8); there was no difference in the mean number of ACE (0·6 in those with and without dementia) (Supplementary Table 4).

In our primary analysis, adulthood adversity score was associated with dementia. Each additional adult adverse experience was associated with a 9% increase in hazard of dementia incidence (95% CI:1·01-1·16). Positive associations were observed between total (adulthood and childhood) adversity scores and dementia incidence however, the 95% CI crossed the null. We did not find evidence to suggest an association between dementia incidence and number of ACEs (Figure 2, Supplementary Table 5). In sensitivity analysis we observed consistent positive associations between adulthood and total adversity scores and dementia incidence. However, the 95% CI crossed the null for both logistic regression models which assumed that everyone censored got dementia. Sensitivity analysis of the association between childhood adversity score and dementia incidence produced estimates close to the null.

**Figure 2.**
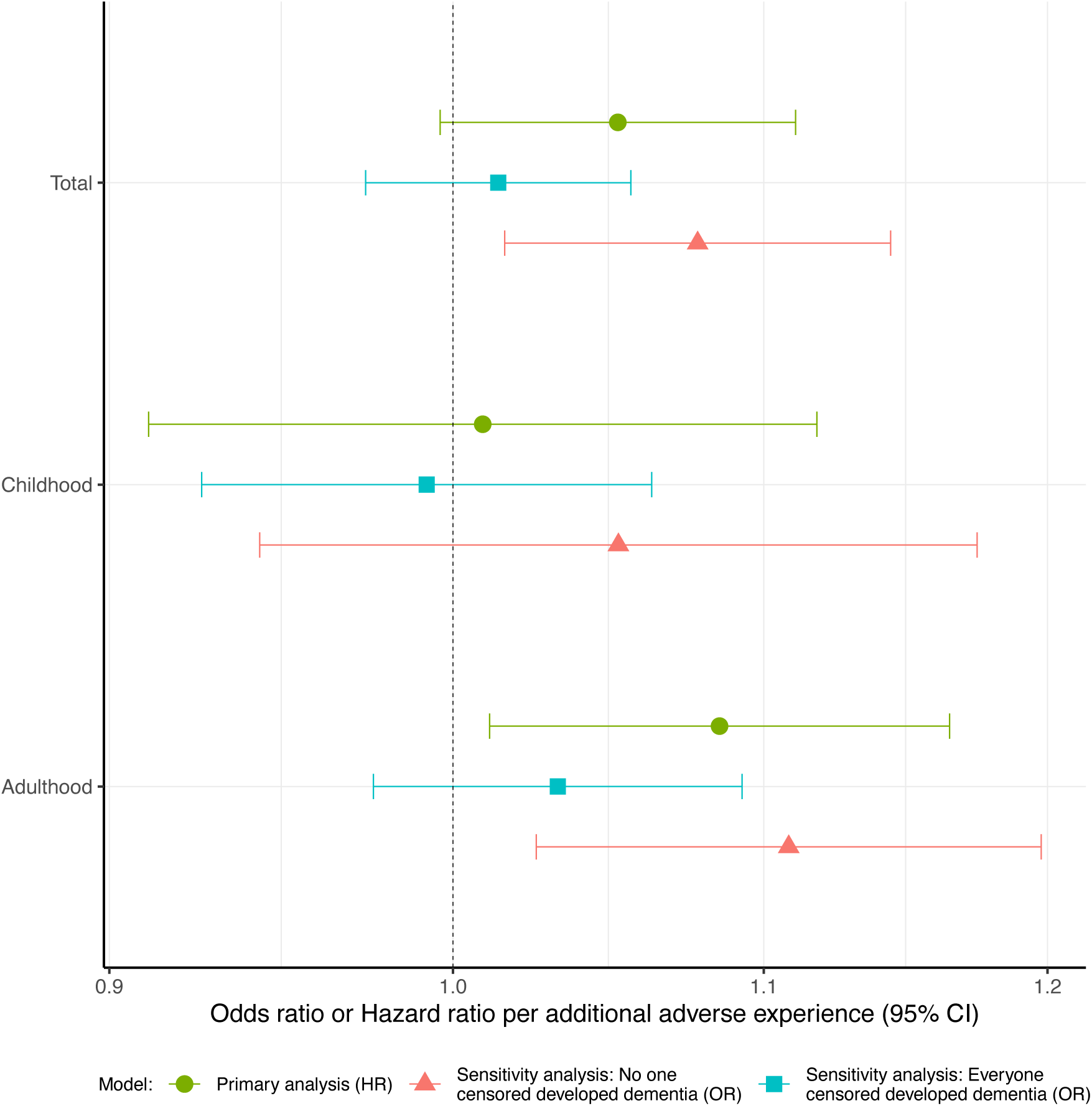
Hazard ratios and odds ratios for dementia incidence per additional adversity experienced. Possible ranges of sum adversity scores are as follows: Total (0-25), Childhood (0-12), Adulthood (0-13). Primary analysis - Cox proportional hazards model of dementia incidence in association with adversity measures, adjusting for sex and ethnicity, using age as the time axis (HR). Sensitivity analysis: No one censored developed dementia – logistic regression of dementia incidence in association with adversity measures, adjusting for age, quadratic age and ethnicity assuming everybody lost to follow up did not develop dementia (OR). Sensitivity analysis: Everyone censored developed dementia – logistic regression of dementia incidence in association with adversity measures, adjusting for age, quadratic age and ethnicity assuming everybody lost to follow up did develop dementia (OR).

When adverse experiences were considered separately, being a victim of a serious physical attack in childhood (HR:2·38, 95% CI:1·18-4·81) or adulthood (HR:1·56, 95% CI:1·05-2·34), having physically abusive parents (HR:2·34, 95% CI:1·56-3·52), experiencing severe financial hardship in adulthood (HR:1·34, 95% CI:1·06-1·68) or having lived in a prisoner of war camp (POW)(HR:6·09, 95% CI:1·49-24·86) were associated with increased dementia risk (Figure 3, Supplementary Table 6). Only five individuals reported being in a POW camp so results should be interpreted with caution. Having spent most of childhood in social care was associated with a 108% increased hazard of developing dementia, although the 95 CI% crossed 1. No adverse experience was protective against dementia.

**Figure 3.**
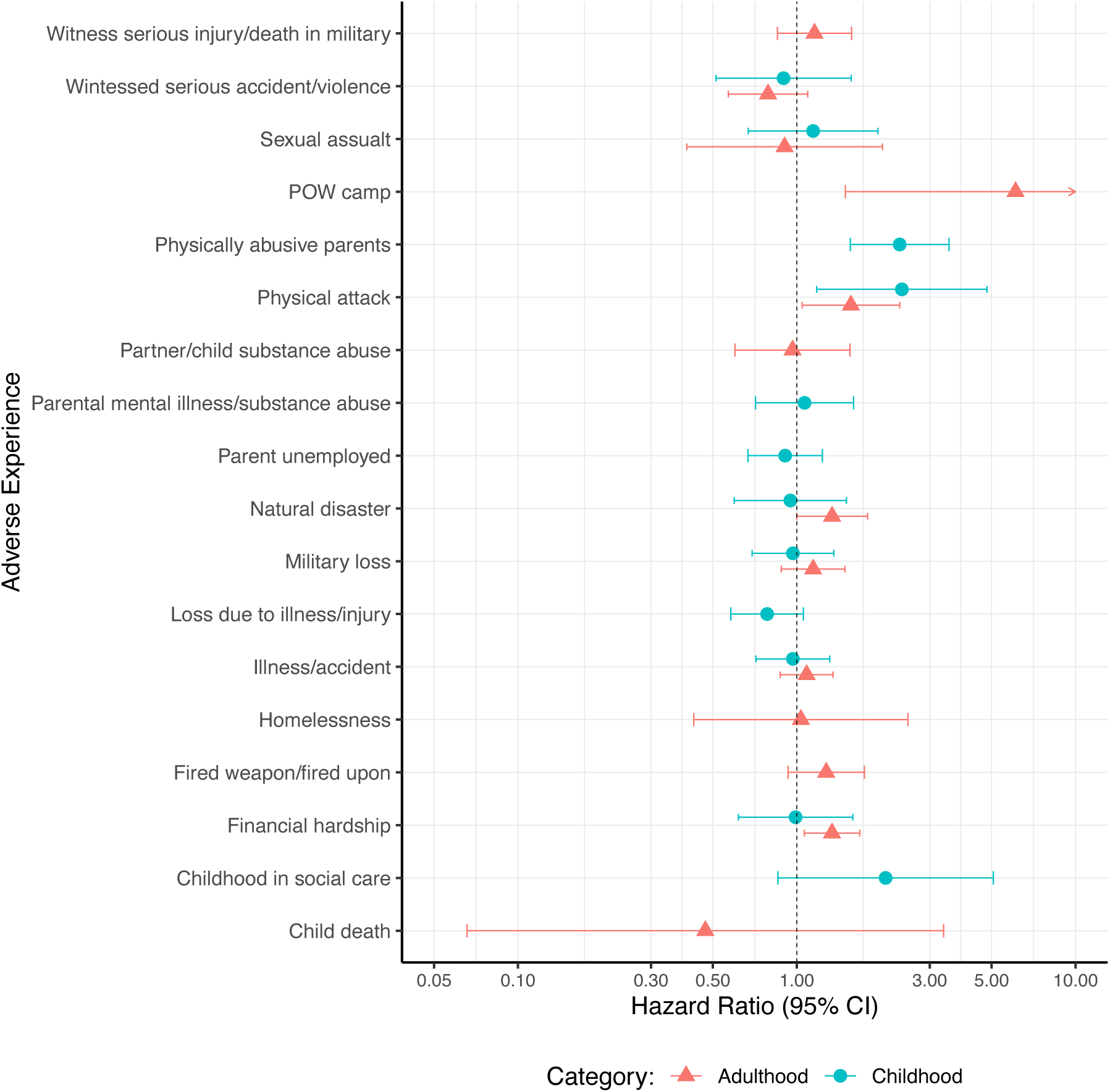
Hazard ratios for dementia according to adverse experiences analysed separately. Cox regression models were adjusted for ethnicity and sex, using age as the time axis.

Individuals who experienced any form of abuse in childhood had a 74% (HR:1·78, 95% CI:1·25-2·43) increased hazard of developing dementia compared to those who did not, adjusting for age, sex, childhood SES and ethnicity (Figure 4, Supplementary Table 7). Associations between dementia and other broad childhood adversities cannot be confidently distinguished from the null. Linear hypothesis testing found evidence of heterogeneity between the HRs (p=0·01, Supplementary Table 7).

**Figure 4.**
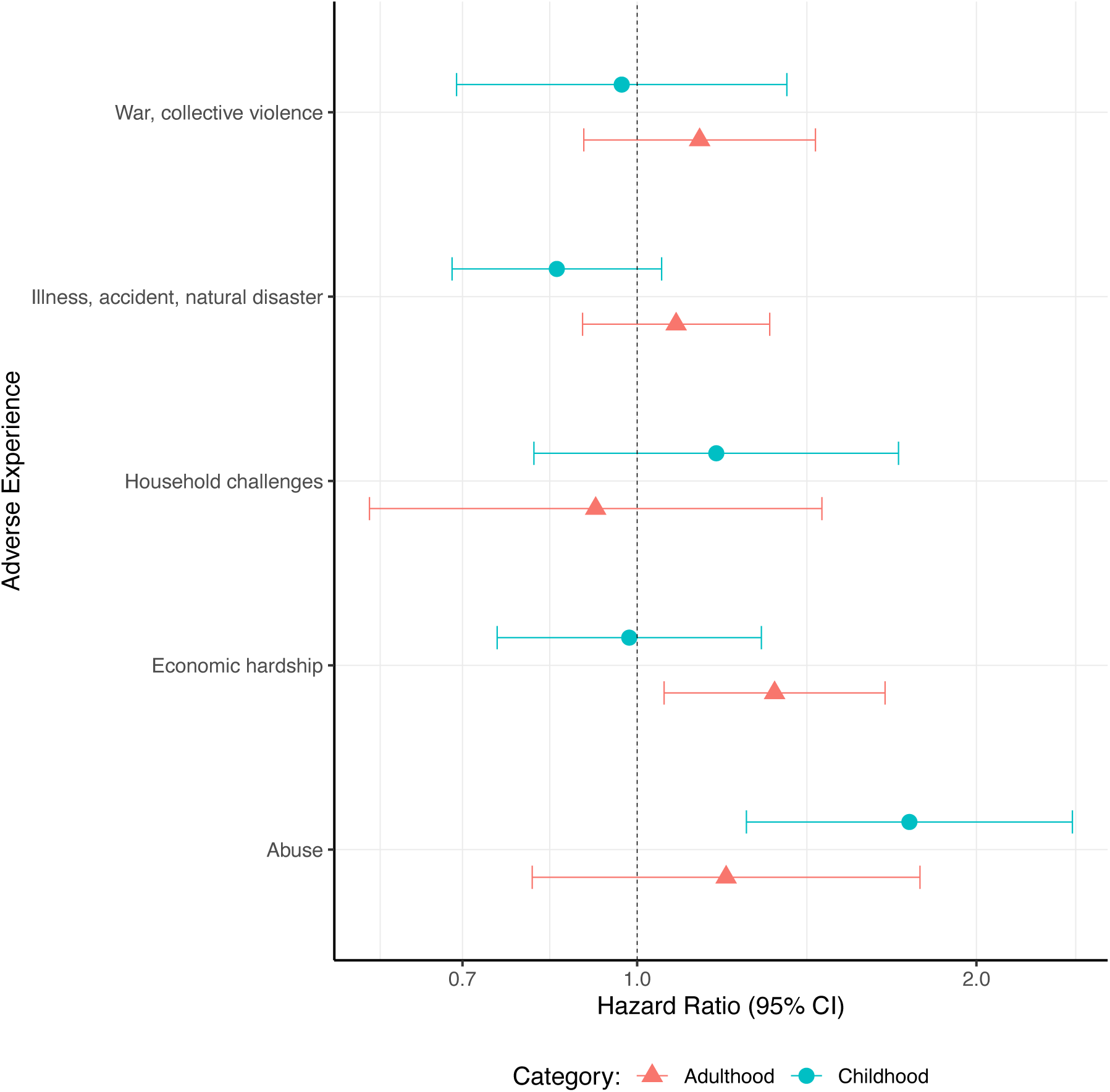
Hazard ratios for dementia incidence in association with broad adversity categories analysed separately. Cox regressions were adjusted for ethnicity, sex and childhood SES, using age as a time axis.

Those who experienced economic hardship in adulthood had a 32% (HR:1·32, 95% CI:1·06-1·66) increased hazard of developing dementia, compared to those who did not, adjusted for age, sex, childhood SES and ethnicity. Positive associations were also observed between dementia incidence and all other broad adult adversity categories aside from household dysfunction, but estimates were imprecise, and 95% CI crossed the null (Figure 4, Supplementary Table 7). We did not find evidence for heterogeneity between the HRs (p=0·49, Supplementary Table 7).

When childhood and adulthood broad categorisations of adversity were included in the same model, there was minimal attenuation of the hazards associated with each adversity, suggesting the hazards are independent of each other. We found some evidence for heterogeneity between the hazards associated with each broad adversity category, but less than when only childhood adversities were considered (p=0·06, Supplementary Table 7).

All estimates investigating effect modification by sex and childhood economic hardship were imprecisely estimated (Supplementary Table 8, Supplementary Table 9). We found limited evidence to suggest associations between adversity measures and dementia differed by sex. Compared to those who did not experience economic hardship in childhood, those who did had higher hazards of dementia in association with all three sum scores of adversities, but the 95% CI overlapped. We found some evidence to suggest that childhood economic hardship modified the relationship between dementia and childhood abuse, adult household challenges and abuse in adulthood, but HR were imprecise.

When analyses were re-run on complete case data, similar findings were observed, though the 95% CI associated with adult economic hardship crossed one (95% CI:0·98-1·64) (Supplementary Table 10, Supplementary Table 11). Sensitivity analysis using categorical measures of sum adversity scores did not reveal a threshold effect (Supplementary Figure 3). No departures from proportional hazards were observed.

## 4. Discussion

### 4.1 Key findings

Within a nationally representative sample of English adults aged over age 50 we observed a linear relationship whereby each additional adverse experience in adulthood was associated with a 9% increased hazard of dementia. Child abuse and adult economic hardship were associated with dementia risk, but little evidence was found for other categories of adverse experiences. We observed significant heterogeneity between the hazards of dementia associated with ACEs, but not adult adverse experiences.

### 4.2 Current literature

Few studies take a cumulative risk approach to investigating the association between adult adverse experiences and dementia. One study found that having at least one serious mid-life stressor was associated with dementia risk, but a dose response relationship was not observed [11]. Frequent and constant psychological stress in mid-life was associated with increased dementia risk in a previous cohort study of females who were asked about psychological stress and followed up for 35 years. However, stressful life events were highly subjective, focussing on perception of stress not an objective experience [12].

We found weak evidence of an association between dementia and total adverse life events across childhood and adulthood. One study found that having three or more traumatic experiences was associated with a 62% increased risk of dementia, compared to having no traumatic experiences [5]. Another observed a dose response relationship between the number of adverse experiences and dementia risk [4]. Our inclusion of high numbers of adverse experiences may have diluted the effect of experiences more strongly associated with dementia.

We did not observe an association between number of ACEs and dementia incidence. Aligning with our findings, a study of Australian older adults analysed ACEs in multiple ways but consistently found null associations [24]. Additionally, within a study of French adults over 65, number of ACEs was not associated with incident dementia [25]. However, multiple studies have observed associations between ACE and dementia [10,14]. Within the UK Biobank, each additional ACE experienced by an individual was associated with a 15·5% increased risk of dementia [14]. However, this study only considers measures of abuse and neglect. Therefore, inconsistencies in the literature could be partially attributed to the consideration of different ACEs, as we found evidence for heterogeneity in their effects.

In line with the sensitive period life-course epidemiology model, several studies observed that stressful experiences in childhood have more potent effects compared to the same experiences if encountered in adulthood [4,6]. Therefore, it is surprising that we find more evidence for associations between dementia and adult adverse experiences. In our study, incidence of childhood adversities was lower than the incidence of adulthood adversities, so estimates relating to ACEs are less precise.

Estimates for adulthood adversity fall within the confidence intervals for childhood adversity estimates, so our results don’t necessarily contradict prior work.

We found that having experienced any type of child abuse was associated with a 74% increased hazard of developing dementia. Consistent with our findings, compared to those with no experience of abuse, individuals who often experienced sexual, physical or emotional abuse had a higher risk of dementia in the UK Biobank [5]. Emotional and sexual abuse, but not physical abuse was associated with dementia in the Panel Study of Income Dynamics. Physical abuse and psychological abuse were associated with dementia incidence in the Japan Gerontological Evaluation Study [9]. However, multiple studies observed null associations between abuse and dementia [24,25]. One study considering Australian adults observed null associations between dementia and verbal abuse, physical abuse and sexual abuse. However, participants were aged 70-79 at the end of follow-up which is lower than the average age of dementia diagnosis [24].

It is well established that low SES is associated with increased dementia risk [26]. Therefore, our finding that adult economic hardship is associated with dementia is expected. In a nationally representative cohort of US older-adults, financial strain was associated with higher odds of incident dementia [27]. Another study observed that resource problems, characterised by financial or employment problems, were associated with 20% greater risk of dementia compared to interpersonal problems [11]. Two studies investigated the association between poverty and dementia but neither concluded there was evidence of an effect [24,25]. However, one evaluated statistical significance at the 99·9% confidence level and found the lower limit was close to the null, suggesting tentative evidence of an effect [24].

It is unexpected that economic hardship in adulthood, but not childhood, is associated with dementia. Given that within our study, economic hardship in adulthood is more prevalent (17·6%) than in childhood (9·2%), parents may shelter children from their economic situation, resulting in an underreporting of childhood economic hardship. Prior work observed that participants who underwent an upwards socioeconomic transition from childhood to adulthood had lower dementia risk, even compared to those who had high SES all their life [28]. Within our study, participants cannot report experiencing severe economic hardship in childhood and adulthood. Therefore, we may be only picking up those on a downwards socioeconomic trajectory. We may underestimate the association between adult economic hardship and dementia by omitting those who experienced economic hardship in childhood and adulthood.

Within this study, sum scores alone are insufficient to understand the impact of adverse life experiences on dementia risk. We find evidence that risk mechanisms differ dependent on whether the experience occurs in childhood or adulthood. When childhood adversity measures are considered, significant heterogeneity with their relation to dementia risk is observed. Additive approaches assume all experiences result in the same impact on dementia risk, but our findings show this assumption is unlikely to hold for ACE. However, by only considering isolated experiences of adversity, we disregard their patterning. Certain traumas are more likely to cluster as shown in our correlation matrixes [8]. Use of person-centred approaches to identify adversities which cluster together could address these limitations in future studies.

These approaches have been used to identify clusters of ACEs, but not adversities encompassing the whole life-course, and are rarely applied to investigate associations with dementia risk [29,30].

### 4.3 Strengths and limitations

ELSA is a well-characterised, nationally representative study, making our findings more generalisable to the older English population than some previous studies. We considered a broad range of adverse experiences across the life-course which enables us to better identify which experiences are associated with dementia. By considering adult adversities alongside childhood adversities, we could examine the importance of the timing of adversity on dementia risk.

Our study lacks statistical power. We aimed to address this by imputing missing data and collating individual adversity measures into broad categories however estimates remained imprecise. Adverse experiences measures were collected retrospectively so could be subject to recall bias; likely biasing results toward the null. Many measures are subjective as we rely on self-reporting of exposures. Individuals may be reluctant to share details of their adverse experiences, resulting in underreporting. Adult adverse experiences are likely to be underreported as the questionnaire design precludes reporting the same experience in both childhood and adulthood.

Therefore, we may have underestimated the association between dementia and adverse experiences. Our study population is mostly White, reducing generalisability to minority groups.

### 4.4 Conclusions

Associations between dementia and adverse experiences throughout the life-course are influenced by the type and timing of events. We find evidence for a cumulative risk effect for adverse experiences in adulthood but observe significant heterogeneity in the impact of ACEs. Taking a more nuanced approach to risk factor measurement and analysis allows us to develop a greater understanding of possible risk mechanisms and to better highlight the importance of acting to reduce the number of people experiencing adverse life events.

## Data Availability

The ELSA data are freely available to researchers through the UK Data Service with access codes SN8346 and 5050.

https://ukdataservice.ac.uk/

## 5 Acknowledgements

This study was supported by the Medical Research Council (Grant MR/W006774/1). The English Longitudinal Study of Ageing is supported by the National Institute on Aging (Grant R01AG017644) and by a consortium of UK Government departments coordinated by the National Institute for Health Research (NIHR).

## 6. Declaration of interests

We declare no competing interests.

## Notes

### Competing Interest Statement

The authors have declared no competing interest.

