## Supplementary Materials for "The association between adverse experiences throughout the life-course and risk of dementia in the English Longitudinal Study of Ageing"

**Supplementary material**

Appendix 1

Most exposure variables were defined as occurring in childhood or adulthood, dependent on the age at which participants reported experiencing the adversity. Some questions answered by participants relate to experiences that occurred prior to the age of 16, so to aid consistency within our analysis we restricted childhood adversities to those which occurred before 16. Where age at adversity was missing due to participants not answering the question “How old were you when it first happened?”, we looked at the proportion of people who had this experience before or after 16. We randomly allocated participants with age at adversity missing to either the childhood adversity or adulthood adversity group, making the proportion in each group the same as when age at adversity data was available. Three variables (had a partner/husband/wife/child addicted to drugs or alcohol; ever fired a weapon in combat or been fired upon; witnessed serious injury/death of someone in war or military combat) were considered unlikely during childhood and so when reported before the age of 16, were treated as missing (N=65). Age sixteen was chosen as the cut off as it is the youngest age that someone can legally join the army, get married, and it is also the legal age of sexual consent in the UK. Individual adversity measures comprise 12 childhood adversity and 13 adult adversity measures, encompassing experiences of household dysfunction, abuse, severe illnesses and military adverse experiences (Table 1).

Many adverse experiences are uncommon, so consolidating similar experiences increased the statistical power of this study. Categories were based on existing literature to consolidate similar adverse experiences. For example, childhood household challenges were defined as having experienced one of the following individual adversity measures: while under the age of 16 parents abused substances or had a mental illness; spent most of childhood in a social care setting. Household challenges and abuse categories were adapted from those described in CDC-Kaiser ACE study (16). The war/collective violence categorisation was guided by the Adverse Childhood Experiences International Questionnaire (17). Similar categorisations were applied to adverse experiences in adulthood and binary measures created to reflect whether an individual had reported any experience within each category.

Appendix 2

At each wave ELSA participants completed the following cognitive tests: number of words correctly recalled after a delay, number of words recalled immediately and knowing today’s date. We considered these measures as they were the only cognitive tests that were consistently completed at each wave. In line with previous work, impairment in each test of cognitive function was defined as 1.5 standard deviations or more below the mean compared with the population aged 50-80 with the same level of education, established using ELSAs baseline cognitive testing data (19). Education level was taken at each wave, and the most recent response was used. Overall impairment was defined as having impairment in at least two domains and scores were deemed invalid if they responded to two tests or fewer. To ensure we weren’t considering transient cognitive declines, we removed transient impairments in cognitive function. If a participant improved by one standard deviation or more in cognitive tests in the consecutive wave, they were considered not to have cognitive impairment. Participants were also asked questions about whether they had any difficulties with activities of daily living (ADLs) at each wave to assess functional impairment. Functional impairment was defined as having difficulties with at least one ADL. If impairment in any ADL was reported once and the participants fully recovered at all further waves of data collection, we considered it to be transient and did not categorise it as functional impairment.

The Informant Questionnaire on Cognitive Decline (IQCODE) is administered where individuals are unable to take cognitive tests which are part of the ELSA protocol at each wave. The IQCODE is made up of 16 questions asking a proxy informant how the participants memory, ability to learn new tasks, judgement or handling of key everyday situations are compared with two years ago. If five or more responses were missing the total score was considered missing. A mean value of the scores from each question was taken and a total score of 3.6 or higher was used to identify cognitive impairment. This threshold has been previously validated through meta-analysis of studies in community setting where both the IQCODE and a validated clinical assessment were carried out to assess for the presence of dementia. This threshold has a high specificity (0.84) and sensitivity (0.82) for dementia compared to ICD-10 code diagnosis (20).

Date of dementia diagnosis was calculated to the nearest month. Where possible, date of dementia diagnosis was taken as reported in wave interviews (N=141). If date of diagnosis was not available, age of dementia diagnosis was used and used to calculate date of diagnosis (N=3). If neither were available, the midpoint between the date of the interview in the wave where dementia was first reported, and their previous interview was used as a proxy (N=371).

Appendix 3

We analysed patterns of missing data to establish whether data were missing at random. We conducted a logistic regression of whether adversity data were missing to observe which covariates acted as predictors of missingness. Data was imputed for 887 participants using chained equations. 20 imputed datasets were generated with a maximum of 10 iterations. Individual adversity measures, age, sex, childhood SES, ethnicity, dementia incidence, time to dementia and censoring by wave 10 were included within the imputation model. Income quintile, education and occupation were included as auxiliary variables. All imputed variables were binary aside from occupation, childhood SES and education which were ordered categorical variables. Sum scores of adversities and broad categorisations of adversity were generated post imputation using the imputed individual adversity measures. The outcome variable had no missing data and therefore follow up time and dementia incidence were not imputed using multiple imputation.

Appendix 4

To investigate the potential for informative censoring to impact our results we conducted logistic regression with dementia as an outcome and total number of adverse experiences, number of childhood adversities and number of adulthood adversities as exposures. Two models for each analysis were estimated, one assuming that no censored participants developed dementia, and one assuming that all censored participants developed dementia. The true association should lie somewhere on a spectrum between the results from the two (22). Age at baseline, sex, childhood SES and ethnicity were adjusted for in each model. Age-squared was also included in all models to account for non-linearity in the relationship between age and dementia incidence.

Supplementary Table 1. Definition of individual adversity measures

| **Individual adverse experiences** | **Date of measurement** | **Questions** | **Broad categorisation** |
| --- | --- | --- | --- |
| **Childhood adversities** | | | |
| Lost a very close friend or relative in war or military combat aged under 16 | Life history interview (February 2007 to August 2007) | Have you ever lost a very close friend or relative in a war or in military service?  How old were you when it first happened? | Exposure to war/collective violence |
| While under the age of 16 parents unemployed for more than 6 months when they wanted to work | Life history interview (February 2007 to August 2007) | When you were aged under 16, were either of your parents unemployed for more than 6 months when they wanted to be working?  How old were you when it first happened? | Economic hardship |
| Experienced severe financial hardship aged under 16 | Life history interview (February 2007 to August 2007) | Have you ever experienced severe financial hardship?  How old were you when it first happened? | Economic hardship |
| Lost a very close friend or relative due to illness or injury aged under 16 | Life history interview (February 2007 to August 2007) | Have you ever had a very close friend or relative who died or was at risk of death due to illness or serious accident?  How old were you when it first happened? | Illness, accidents and natural disasters |
| Witnessed an accident/violence which caused death or serious injury (not war) aged under 16 | Life history interview (February 2007 to August 2007) | Other than in war or military action, have you ever witnessed an accident or violent act in which someone was killed or seriously wounded?  How old were you when it first happened? | Illness, accidents and natural disasters |
| Experienced a natural disaster aged under 16 | Life history interview (February 2007 to August 2007) | Have you ever experienced a major fire, flood, earthquake or other natural disaster?  How old were you when it first happened? | Illness, accidents and natural disasters |
| Had a life-threatening illness or accident aged under 16 | Life history interview (February 2007 to August 2007) | Have you ever had a life-threatening illness or accident?  How old were you when it first happened? | Illness, accidents and natural disasters |
| While under the age of 16 parents abused substances or had a mental illness | Life history interview (February 2007 to August 2007) | When you were aged under 16, did your parents drink excessively, take drugs or have mental health problems?  How old were you when it first happened? | Household challenges |
| Spent most of childhood in a social care setting | Wave 1 (March 2002–March 2003)  Wave 2 (June 2004–June 2005)  Wave 3 (May 2006–August 2007) | Who did you live with for most of your childhood?  Foster Parents  Children's Home | Household challenges |
| Sexual assault aged under 16 | Life history interview (February 2007 to August 2007) | Have you ever been a victim of sexual assault (including rape or harassment)?  How old were you when it first happened? | Abuse |
| Physically abusive parents aged 16 or under | Life history interview (February 2007 to August 2007) | When you were aged under 16, were you physically abused by your parents?  How old were you when it first happened? | Abuse |
| Victim of serious physical attack aged under 16 | Life history interview (February 2007 to August 2007) | Have you ever been a victim of serious physical attack or assault?  How old were you when it first happened? | Abuse |
| **Adult adversities** | | | |
| Lost a very close friend or relative in war or military combat aged 16 or over | Life history interview (February 2007 to August 2007) | Have you ever lost a very close friend or relative in a war or in military service?  How old were you when it first happened? | Exposure to war/collective violence |
| Ever fired a weapon in combat or be fired upon | Life history interview (February 2007 to August 2007) | Have you ever fired a weapon in combat or been fired upon?  How old were you when it first happened? | Exposure to war/collective violence |
| Witnessed serious injury/death of someone in war or military combat | Life history interview (February 2007 to August 2007) | Have you ever witnessed the serious injury or death of someone in war or military action?  How old were you when it first happened? | Exposure to war/collective violence |
| Have lived in a prisoner of war camp | Life history interview (February 2007 to August 2007) | Can I check, have you ever experienced any of the things on this card?  Lived in a prisoner of war camp | Exposure to war/collective violence |
| Experienced severe financial hardship aged 16 or over | Life history interview (February 2007 to August 2007) | Have you ever experienced severe financial hardship?  How old were you when it first happened? | Economic hardship |
| Have been homeless for one month or more | Life history interview (February 2007 to August 2007) | Can I check, have you ever experienced any of the things on this card?  Been homeless for 1 month of more? | Economic hardship |
| Lost a very close friend or relative due to illness or injury aged 16 or over | Life history interview (February 2007 to August 2007) | Have you ever had a very close friend or relative who died or was at risk of death due to illness or serious accident?  How old were you when it first happened? | Illness, accidents and natural disasters |
| Witnessed an accident/violence which caused death or serious injury (not war) aged 16 or over | Life history interview (February 2007 to August 2007) | Other than in war or military action, have you ever witnessed an accident or violent act in which someone was killed or seriously wounded?  How old were you when it first happened? | Illness, accidents and natural disasters |
| Had a life-threatening illness or accident aged 16 or over | Life history interview (February 2007 to August 2007) | Have you ever had a life-threatening illness or accident?  How old were you when it first happened? | Illness, accidents and natural disasters |
| Experienced a natural disaster aged 16 or over | Life history interview (February 2007 to August 2007) | Have you ever experienced a major fire, flood, earthquake or other natural disaster?  How old were you when it first happened? | Illness, accidents and natural disasters |
| Have a child who has died | Life history interview (February 2007 to August 2007) | Our records show that when we last interviewed you, you had a child called [^Name of child], whose date of birth was [^Child’s date of birth]. Are these details correct?  Is this child still alive? | Household challenges |
| Had a partner/husband/wife/child addicted to drugs or alcohol | Life history interview (February 2007 to August 2007) | Have you ever had a husband, wife, partner or child who has been addicted to drugs or alcohol?  How old were you when it first happened? | Household challenges |
| Sexual assault aged 16 or over | Life history interview (February 2007 to August 2007) | Have you ever been a victim of sexual assault (including rape or harassment)?  How old were you when it first happened? | Abuse |
| Victim of serious physical attack aged 16 or over | Life history interview (February 2007 to August 2007) | Have you ever been a victim of serious physical attack or assault?  How old were you when it first happened? | Abuse |

Supplementary Table 2. Sample characteristics of those included and excluded in our study.

|  | **Analytical Sample (N=5450)** | **Excluded (N=3360)** |
| --- | --- | --- |
| **Age at wave 3** |  |  |
| Mean (SD) | 65.4 (9.8) | 66.8 (12.4) |
| **Income quintile** |  |  |
| 1 | 800 (15.1%) | 781 (24.7%) |
| 2 | 941 (17.7%) | 716 (22.6%) |
| 3 | 1119 (21.1%) | 595 (18.8%) |
| 4 | 1163 (21.9%) | 561 (17.7%) |
| 5 | 1282 (24.2%) | 513 (16.2%) |
| Missing | 145 (2.7%) | 194 (5.8%) |
| **Sex** |  |  |
| Male | 2397 (44.0%) | 1544 (46.0%) |
| Female | 3053 (56.0%) | 1816 (54.0%) |
| Missing | 0 (0.0%) | 0 (0.0%) |
| **Ethnicity** |  |  |
| White | 5362 (98.4%) | 3207 (95.5%) |
| Other ethnicities | 88 (1.6%) | 152 (4.5%) |
| Missing | 0 (0.0%) | 1 (0.0%) |
| **Occupation** |  |  |
| Managerial and professional occupations | 1846 (33.9%) | 956 (28.5%) |
| Intermediate occupations | 1420 (26.1%) | 766 (22.9%) |
| Routine and manual occupations | 2124 (39.0%) | 1558 (46.5%) |
| Other | 59 (1.1%) | 70 (2.1%) |
| Missing | 1 (0.00%) | 10 (0.3%) |
| **Education category** |  |  |
| No qualification | 1947 (35.7%) | 1494 (44.5%) |
| High school level | 1681 (30.8%) | 956 (28.5%) |
| University level | 1821 (33.4%) | 908 (27.0%) |
| Missing | 1 (0.0%) | 2 (0.1%) |
| **Dementia incidence during follow-up** |  |  |
| No | 4935 (90.6%) | 3059 (91.0%) |
| Yes | 515 (9.4%) | 301 (9.0%) |
| Missing | 0 (0.0%) | 0 (0.0%) |

Supplementary Table 3. Sample characteristics of complete case and imputed data.

|  | **Complete case data** | | **Imputed data** |
| --- | --- | --- | --- |
| **Variable** | **Categorical variables: N (%) / Continuous variables: N (SD)** | **Missing (%)** | **Categorical variables: proportion / Continuous variables: mean (SD)** |
| **Age**  Mean (SD) | 66.1 (9.8) | 0 (0.0%) | 66.1 (9.8) |
| **Income quintile**  1 2 3 4 5 | 800 (15.1%) 941 (17.7%) 1119 (21.1%) 1163 (21.9%) 1282 (24.2%) | 145 (2.7%) | 15.0% 17.7% 21.1% 21.9% 24.3% |
| **Sex**  Male Female | 2397 (44.0%) 3053 (56.0%) | 0 (0.0%) | 44.0% 56.0% |
| **Ethnicity**  White Other ethnicities | 5362 (98.4%) 88 (1.6%) | 0 (0.0%) | 98.4% 1.6% |
| **Occupation**  Managerial and professional occupations Intermediate occupations Routine and manual occupations Other | 1846 (33.9%) 1420 (26.1%) 2124 (39.0%) 59 (1.1%) | 1 (0.0%) | 33.9% 26.1% 39.0% 1.1% |
| **Education category**  No qualification High school level University level | 1947 (35.7%) 1681 (30.8%) 1821 (33.4%) | 1 (0.0%) | 35.7% 30.8% 33.4% |
| **Number of books in the house at age 10**  More than 10 books  10 books or fewer | 3851 (74.0%) 1356 (26.0%) | 243 (4.5%) | 73.8% 26.2% |
| **Time under follow-up (months)**  Mean (sd) | 121.4 (57.3) | 0 (0.0%) | 121.4 (57.3) |
| **Dementia incidence during follow up**  No Yes | 4935 (90.6%) 515 (9.4%) | 0 (0.0%) | 90.6% 9.4% |
| **Adverse childhood experiences** | | | |
| **Spent most of childhood in a social care setting**  No Yes | 5420 (99.4%) 30 (0.6%) | 0 (0.0%) | 99.4% 0.6% |
| **Sexual assault aged under 16**  No Yes | 5194 (96.0%) 214 (4.0%) | 42 (0.8%) | 96% 4% |
| **Victim of serious physical attack aged under 16**  No Yes | 5330 (98.6%) 77 (1.4%) | 43 (0.8%) | 98.6% 1.4% |
| **Physically abusive parents aged 16 or under**  No Yes | 5226 (96.6%) 183 (3.4%) | 41 (0.8%) | 96.6% 3.4% |
| **While under the age of 16 parents abused substances or had a mental illness**  No Yes | 5076 (94.1%) 318 (5.9%) | 56 (1.0%) | 94.1% 5.9% |
| **Experienced a natural disaster aged under 16**  No Yes | 5221 (96.6%) 185 (3.4%) | 44 (0.8%) | 96.5% 3.5% |
| **While under the age of 16 parents unemployed for more than 6 months when they wanted to work**  No Yes | 4996 (93.0%) 378 (7.0%) | 76 (1.4%) | 92.9% 7.1% |
| **Had a life-threatening illness or accident aged under 16**  No Yes | 4924 (91.2%) 474 (8.8%) | 52 (1.0%) | 91.2% 8.8% |
| **Lost a very close friend or relative in war of military combat aged under 16**  No Yes | 5135 (95.2%) 257 (4.8%) | 58 (1.1%) | 95.2% 4.8% |
| **Lost a very close friend or relative due to illness or injury aged under 16**  No Yes | 4665 (87.3%) 679 (12.7%) | 106 (1.9%) | 87.3% 12.7% |
| **Witnessed an accident/violence which caused death or serious injury (not war) aged under 16**  No Yes | 5220 (96.9%) 169 (3.1%) | 61 (1.1%) | 96.9% 3.1% |
| **Experienced severe financial hardship aged under 16**  No Yes | 5237 (97.2%) 153 (2.8%) | 60 (1.1%) | 97.1% 2.9% |
| **Adverse adult experiences** | | | |
| **Sexual assault aged 16 or over**  No Yes | 5294 (97.9%) 114 (2.1%) | 42 (0.8%) | 97.9% 2.1% |
| **Victim of serious physical attack aged 16 or over**  No Yes | 5169 (95.6%) 238 (4.4%) | 43 (0.8%) | 95.6% 4.4% |
| **Experienced a natural disaster aged 16 or over**  No Yes | 5020 (92.9%) 386 (7.1%) | 44 (0.8%) | 92.8% 7.2% |
| **Have a child who has died**  No Yes | 5411 (99.7%) 15 (0.3%) | 24 (0.4%) | 99.7% 0.3% |
| **Have lived in a prisoner of war camp**  No Yes | 5442 (99.9%) 5 (0.1%) | 3 (0.1%) | 99.9% 0.1% |
| **Have been homeless for one month or more**  No Yes | 5374 (98.7%) 73 (1.3%) | 3 (0.1%) | 98.7% 1.3% |
| **Had a life-threatening illness or accident aged 16 or over**  No Yes | 4452 (82.5%) 946 (17.5%) | 52 (1.0%) | 82.4% 17.6% |
| **Had a partner/husband/wife/child addicted to drugs or alcohol**  No Yes | 5154 (95.2%) 262 (4.8%) | 34 (0.6%) | 95.1% 4.9% |
| **Ever fired a weapon in combat or be fired upon**  No Yes | 5128 (94.9%) 274 (5.1%) | 48 (0.9%) | 94.9% 5.1% |
| **Witness serious injury/death of someone in war or military combat**  No Yes | 5105 (94.9%) 273 (5.1%) | 72 (1.3%) | 94.8% 5.2% |
| **Lost a very close friend or relative in war of military combat aged 16 or over**  No Yes | 5023 (93.2%) 369 (6.8%) | 58 (1.1%) | 93.1% 6.9% |
| **Witnessed an accident/violence which caused death or serious injury (not war) aged 16 or over**  No Yes | 4810 (89.3%) 579 (10.7%) | 61 (1.1%) | 89.2% 10.8% |
| **Experienced severe financial hardship aged 16 or over**  No Yes | 4484 (83.2%) 906 (16.8%) | 60 (1.1%) | 83.2% 16.8% |
| **Sum adversity score** | | | |
| **Total number of adversities**  Mean (sd) | 1.4 (1.5) | 558 (10.2%) | 1.4 (1.5) |
| **Number of adult adversities**  Mean (sd) | 0.8 (1.1) | 477 (8.8%) | 0.8 (1.1) |
| **Number of childhood adversities**  Mean (sd) | 0.6 (0.9) | 443 (8.1%) | 0.6 (0.9) |
| **Broad adversity measures** | | | |
| **Childhood exposure to war/collective violence**  No Yes | 5135 (95.2%) 257 (4.8%) | 58 (1.1%) | 95.2% 4.8% |
| **Childhood economic hardship**  No Yes | 4838 (90.9%) 483 (9.1%) | 129 (2.4%) | 90.8% 9.2% |
| **Childhood exposure to illness, accident or natural disaster**  No Yes | 3956 (75.7%) 1270 (24.3%) | 224 (4.1%) | 75.8% 24.2% |
| **Childhood household challenges**  No Yes | 5059 (93.8%) 335 (6.2%) | 56 (1.0%) | 93.6% 6.4% |
| **Childhood abuse**  No Yes | 4959 (92.7%) 389 (7.3%) | 102 (1.9%) | 92.5% 7.5% |
| **Adult exposure to war/collective violence**  No Yes | 4737 (89.4%) 564 (10.6%) | 149 (2.7%) | 89.0% 10.9% |
| **Adult economic hardship**  No Yes | 4443 (82.5%) 944 (17.5%) | 63 (1.2%) | 82.4% 17.6% |
| **Adult exposure to illness, accident or natural disaster**  No Yes | 3772 (71.0%) 1539 (29.0%) | 139 (2.6%) | 70.7% 29.3% |
| **Adult household challenges**  No Yes | 5118 (94.9%) 274 (5.1%) | 58 (1.1%) | 94.9% 5.1% |
| **Adult childhood abuse**  No Yes | 5062 (94.1%) 318 (5.9%) | 70(1.3%) | 94.0% 6.0% |

Details of the imputation procedure are given in Appendix 3.

Supplementary Table 4. Descriptive analysis of covariates by dementia incidence.

| **Variable** | **Dementia**  **(9.4%)** | **No dementia**  **(80.6%)** |
| --- | --- | --- |
| **Age at life history interview**  Mean (sd) | 74.0 (8.8) | 65.3 (9.5) |
| **Income quintile**  1 2 3 4 5 | 19.0% 19.0% 22.0% 20.0% 20.0% | 15.0% 17.0% 21.0% 22.0% 25.0% |
| **Sex**  Male Female | 46.0% 54.0% | 44.0% 56.0% |
| **Ethnicity**  White Other ethnicities | 98.0% 2.0% | 98.0% 2.0% |
| **Occupation**  Managerial and professional occupations Intermediate occupations Routine and manual occupations Other | 31.0% 26.0% 42.0% 2.0% | 34.0% 26.0% 39.0% 1.0% |
| **Education category**  No qualification High school level University level | 42.0% 31.0% 27.0% | 35.0% 31.0% 34.0% |
| **Number of books in the house at age 10**  More than 10 books  10 books or fewer | 64.0% 38.0% | 75.0% 25.0% |
| **Time under follow-up (months)**  Mean (sd) | 78.8 (48.7) | 125.8 (56.34) |
| **Adverse childhood experiences** | | |
| **Spent most of childhood in a social care setting**  No Yes | 99.00% 1.0% | 99.0% 1.0% |
| **Sexual assault aged under 16**  No Yes | 97.0% 3.0% | 96.0% 4.0% |
| **Victim of serious physical attack aged under 16**  No Yes | 98.0% 2.0% | 99.0% 1.0% |
| **Physically abusive parents aged 16 or under**  No Yes | 95.0% 5.0% | 97.0% 3.0% |
| **While under the age of 16 parents abused substances or had a mental illness**  No Yes | 95.0% 5.0% | 94.0% 6.0% |
| **Experienced a natural disaster aged under 16**  No Yes | 96.0% 4.0% | 97.0% 3.0% |
| **While under the age of 16 parents unemployed for more than 6 months when they wanted to work**  No Yes | 91.0% 9.0% | 93.0% 7.0% |
| **Had a life-threatening illness or accident aged under 16**  No Yes | 91.0% 9.0% | 91.0% 9.0% |
| **Lost a very close friend or relative in war of military combat aged under 16**  No Yes | 93.0% 7.0% | 95.0% 5.0% |
| **Lost a very close friend or relative due to illness or injury aged under 16**  No Yes | 90.0% 10.0% | 87.0% 13.0% |
| **Witnessed an accident/violence which caused death or serious injury (not war) aged under 16**  No Yes | 97.0% 3.0% | 97.0% 3.0% |
| **Experienced severe financial hardship aged under 16**  No Yes | 96.0% 4.0% | 97.0% 3.0% |
| **Adverse adult experiences** | | |
| **Sexual assault aged 16 or over**  No Yes | 99.0% 1.0% | 98.0% 2.0% |
| **Victim of serious physical attack aged 16 or over**  No Yes | 95.0% 5.0% | 96.0% 4.0% |
| **Experienced a natural disaster aged 16 or over**  No Yes | 90.0% 10.0% | 93.0% 7.0% |
| **Have a child who has died**  No Yes | 100.0% 0.0% | 100.0% 0.0% |
| **Have lived in a prisoner of war camp**  No Yes | 100.0% 0.0% | 100.0% 0.0% |
| **Have been homeless for one month or more**  No Yes | 99.0% 1.0% | 99.0% 1.0% |
| **Had a life-threatening illness or accident aged 16 or over**  No Yes | 80.0% 20.0% | 83.0% 17.0% |
| **Had a partner/husband/wife/child addicted to drugs or alcohol**  No Yes | 96.0% 4.0% | 95.0% 5.0% |
| **Ever fired a weapon in combat or be fired upon**  No Yes | 90.0% 10.0% | 95.0% 5.0% |
| **Witness serious injury/death of someone in war or military combat**  No Yes | 90.0% 10.0% | 95.0% 5.0% |
| **Lost a very close friend or relative in war of military combat aged 16 or over**  No Yes | 85.0% 15.0% | 94.0% 6.0% |
| **Witnessed an accident/violence which caused death or serious injury (not war) aged 16 or over**  No Yes | 92.0% 8.0% | 89.0% 11.0% |
| **Experienced severe financial hardship aged 16 or over**  No Yes | 82.0% 18.0% | 83.0% 17.0% |
| **Sum adversity score** | | |
| **Total number of adversities**  Mean (sd) | 1.6 (1.8) | 1.4 (1.5) |
| **Number of adult adversities**  Mean (sd) | 1.0 (1.3) | 0.8 (1.1) |
| **Number of childhood adversities**  Mean (sd) | 0.6 (0.9) | 0.6 (0.9) |
| **Broad adversity measures** | | |
| **Childhood exposure to war/collective violence**  No Yes | 93.0% 7.0% | 95.0% 5.0% |
| **Childhood economic hardship**  No Yes | 88.0% 12.0% | 91.0% 9.0% |
| **Childhood exposure to illness, accident or natural disaster**  No Yes | 78.0% 22.0% | 75.0% 25.0% |
| **Childhood household challenges**  No Yes | 94.0% 6.0% | 94.0% 6.0% |
| **Childhood abuse**  No Yes | 92.0% 8.0% | 93.0% 7.0% |
| **Adult exposure to war/collective violence**  No Yes | 79.0% 21.0% | 90.0% 10.0% |
| **Adult economic hardship**  No Yes | 81.0% 19.0% | 83.0% 17.0% |
| **Adult exposure to illness, accident or natural disaster**  No Yes | 68.0% 32.0% | 71.0% 29.0% |
| **Adult household challenges**  No Yes | 96.0% 4.0% | 95.0% 5.0% |
| **Adult childhood abuse**  No Yes | 95.0% 5.0% | 94.0% 6.0% |

Supplementary Table 5. Hazard ratios and odds ratios for dementia incidence per additional adversity experienced.

| **Total number of adversities** | | | |
| --- | --- | --- | --- |
| **Primary Analysis** | **HR** | **95% CI** | |
| Cox proportional hazards model | 1.05 | 1.00 | 1.11 |
| **Sensitivity Analysis** | **OR** | **95% CI** | |
| Logistic regression: no one censored developed dementia | 1.08 | 1.02 | 1.14 |
| Logistics regression: everyone censored developed dementia | 1.01 | 0.97 | 1.06 |
| **Number of adult adversities** | | | |
| **Primary Analysis** | **HR** | **95% CI** | |
| Cox proportional hazards model | 1.09 | 1.01 | 1.16 |
| **Sensitivity Analysis** | **OR** | **95% CI** | |
| Logistic regression: no one censored developed dementia | 1.11 | 1.03 | 1.20 |
| Logistics regression: everyone censored developed dementia | 1.03 | 0.98 | 1.09 |
| **Number of childhood adversities** | | | |
| **Primary Analysis** | **HR** | **95% CI** | |
| Cox proportional hazards model | 1.01 | 0.91 | 1.12 |
| **Sensitivity Analysis** | **OR** | **95% CI** | |
| Logistic regression: no one censored developed dementia | 1.05 | 0.94 | 1.17 |
| Logistics regression: everyone censored developed dementia | 0.99 | 0.93 | 1.06 |

Possible ranges of sum adversity scores are as follows: Total (0-25), Childhood (0-12), Adulthood (0-13). Primary analysis - Cox proportional hazards model of dementia incidence in association with adversity measures, adjusting for sex and ethnicity, using age as the time axis (HR). Sensitivity analysis: No one censored developed dementia – logistic regression of dementia incidence in association with adversity measures, adjusting for age, quadratic age and ethnicity assuming everybody lost to follow up did not develop dementia (OR). Sensitivity analysis: Everyone censored developed dementia – logistic regression of dementia incidence in association with adversity measures, adjusting for age, quadratic age and ethnicity assuming everybody lost to follow up did develop dementia (OR).

Supplementary Table 6. Cox regression of dementia in association with adverse experiences analysed separately, adjusted for ethnicity and sex and using age as the time axis

| **Cox regression** | **HR** | **95% CI** | |
| --- | --- | --- | --- |
| **Adverse childhood experiences** | | | |
| Spent most of childhood in a social care setting | 2.08 | 0.86 | 5.07 |
| Sexual assault aged under 16 | 1.14 | 0.67 | 1.95 |
| Victim of serious physical attack aged under 16 | 2.38 | 1.18 | 4.81 |
| Physically abusive parents aged 16 or under | 2.34 | 1.56 | 3.52 |
| While under the age of 16 parents abused substances or had a mental illness | 1.07 | 0.71 | 1.60 |
| Experienced a natural disaster aged under 16 | 0.95 | 0.60 | 1.51 |
| While under the age of 16 parents unemployed for more than 6 months when they wanted to work | 0.91 | 0.67 | 1.24 |
| Had a life-threatening illness or accident aged under 16 | 0.97 | 0.71 | 1.31 |
| Lost a very close friend or relative in war of military combat aged under 16 | 0.97 | 0.69 | 1.36 |
| Lost a very close friend or relative due to illness or injury aged under 16 | 0.78 | 0.58 | 1.06 |
| Witnessed an accident/violence which caused death or serious injury (not war) aged under 16 | 0.90 | 0.51 | 1.57 |
| Experienced severe financial hardship aged under 16 | 0.99 | 0.62 | 1.59 |
| **Adverse adult experiences** | | | |
| Sexual assault aged 16 or over | 0.90 | 0.40 | 2.03 |
| Victim of serious physical attack aged 16 or over | 1.56 | 1.05 | 2.34 |
| Experienced a natural disaster aged 16 or over | 1.34 | 1.00 | 1.79 |
| Have a child who has died | 0.47 | 0.07 | 3.36 |
| Have lived in a prisoner of war camp | 6.09 | 1.49 | 24.86 |
| Have been homeless for one month or more | 1.03 | 0.43 | 2.51 |
| Had a life-threatening illness or accident aged 16 or over | 1.08 | 0.87 | 1.35 |
| Had a partner/husband/wife/child addicted to drugs or alcohol | 0.97 | 0.60 | 1.55 |
| Ever fired a weapon in combat or be fired upon | 1.28 | 0.93 | 1.75 |
| Witness serious injury/death of someone in war or military combat | 1.16 | 0.85 | 1.57 |
| Lost a very close friend or relative in war of military combat aged 16 or over | 1.14 | 0.88 | 1.49 |
| Witnessed an accident/violence which caused death or serious injury (not war) aged 16 or over | 0.79 | 0.57 | 1.09 |
| Experienced severe financial hardship aged 16 or over | 1.34 | 1.06 | 1.68 |

Supplementary Table 7. Hazard rations for dementia in association with broad adversity categories. Broad categories were analysed individually and then mutually adjusted for each other. Cox models were also adjusted for sex, ethnicity and childhood SES. Age was used as the time axis.

|  | **Individual analysis** | **Mutually adjusted** |
| --- | --- | --- |
| **Adverse experience** | **HR (95% CI)** | **HR (95% CI)** |
| **Childhood adversities** |  |  |
| Exposure to war/collective violence | 0.97 (0.69-1.36) | 0.96 (0.69-1.35) |
| Economic hardship | 0.98 (0.75-1.29) | 1.00 (0.76-1.31) |
| Abuse | 1.74 (1.25-2.43) | 1.72 (1.22-2.40) |
| Illness, accident and natural disaster | 0.85 (0.69-1.05) | 0.85 (0.68-1.05) |
| Household challenges | 1.18 (0.81-1.71) | 1.09 (0.74-1.59) |
| Wald test p-value |  | 0.01 |
| **Adult adversities** |  |  |
| Exposure to war/collective violence | 1.14 (0.90-1.44) | 1.16 (0.92-1.46) |
| Economic hardship | 1.32 (1.06-1.66) | 1.31 (1.04-1.65) |
| Abuse | 1.20 (0.81-1.78) | 1.14 (0.76-1.70) |
| Illness, accident and natural disaster | 1.08 (0.89-1.31) | 1.07 (0.89-1.30) |
| Household challenges | 0.92 (0.58-1.46) | 0.80 (0.50-1.29) |
| Wald test p-value |  | 0.49 |
| **Adult and childhood adversities** |  |  |
| Childhood exposure to war/collective violence | 0.97 (0.69-1.36 | 0.98 (0.70-1.38) |
| Childhood economic hardship | 0.98 (0.75-1.29) | 0.98 (0.74-1.29) |
| Childhood abuse | 1.74 (1.25-2.43) | 1.65 (1.17-2.33) |
| Childhood illness, accident and natural disaster | 0.85 (0.69-1.05) | 0.83 (0.67-1.03) |
| Childhood household challenges | 1.18 (0.81-1.71) | 1.04 (0.71-1.53) |
| Adult exposure to war/collective violence | 1.14 (0.90-1.44) | 1.15 (0.92-1.45) |
| Adult economic hardship | 1.32 (1.06-1.66) | 1.29 (1.02-1.63) |
| Adult abuse | 1.20 (0.81-1.78) | 1.09 (0.73-1.64) |
| Adult illness, accident and natural disaster | 1.08 (0.89-1.31) | 1.07 (0.88-1.29) |
| Adult household challenges | 0.92 (0.58-1.46) | 0.76 (0.47-1.22) |
| Wald test p-value |  | 0.06 |

Supplementary Table 8. Interaction between adverse experiences and sex. Cox models were adjusted for ethnicity and childhood SES. Age was used as the time axis.

|  | **Male** | **Female** |
| --- | --- | --- |
| **Cox regression** | **HR (95% CI)** | **HR (95% CI)** |
| **Cumulative measures** | | |
| Total number of adversities | 1.05 (0.98-1.12) | 1.06 (0.93-1.20) |
| Number of adult adversities | 1.07 (0.99-1.17) | 1.11 (0.93-1.31) |
| Number of childhood adversities | 1.01 (0.87-1.16) | 1.01 (0.79-1.30) |
| **Number of childhood adversities** | | |
| Exposure to war/collective violence | 1.20 (0.75-1.92) | 0.71 (0.44-1.16) |
| Economic hardship | 0.98 (0.67-1.43) | 0.85 (0.58-1.25) |
| Illness, accident and natural disaster | 0.91 (0.67-1.22) | 0.70 (0.51-0.96) |
| Household challenges | 0.97 (0.53-1.78) | 1.13 (0.71-1.81) |
| Abuse | 1.71 (0.99-2.95) | 1.50 (0.98-2.27) |
| **Number of adulthood adversities** | | |
| Exposure to war/collective violence | 1.16 (0.87-1.55) | 0.97 (0.66-1.44) |
| Economic hardship | 1.23 (0.86-1.76) | 1.16 (0.87-1.55) |
| Illness, accident and natural disaster | 1.03 (0.80-1.34) | 0.97 (0.72-1.29) |
| Household challenges | 1.44 (0.71-2.92) | 0.64 (0.35-1.17) |
| Abuse | 1.39 (0.81-2.38) | 0.91 (0.51-1.63) |

Supplementary Table 9. Interaction between adverse experiences and childhood economic hardship. Cox models were adjusted for sex, ethnicity and childhood SES. Age was used as the time axis.

|  | **No childhood economic hardship** | **Childhood economic hardship** |
| --- | --- | --- |
| **Cox regression** | **HR (95% CI)** | **HR (95% CI)** |
| **Sum adversity score** | | |
| Total number of adversities | 1.05 (0.98-1.12) | 1.15 (0.97-1.35) |
| Number of adult adversities | 1.07 (1.00-1.16) | 1.16 (0.94-1.43) |
| Number of childhood adversities | 1.00 (0.87-1.14) | 1.17 (0.85-1.59) |
| **Broad childhood trauma measures** | | |
| Exposure to war/collective violence | 0.96 (0.67-1.39) | 1.00 (0.44-2.24) |
| Illness, accident and natural disaster | 0.83 (0.65-1.05) | 0.93 (0.59-1.44) |
| Household challenges | 1.18 (0.76-1.82) | 1.16 (0.57-2.34) |
| Abuse | 1.65 (1.14-2.38) | 2.21 (1.05-4.67) |
| **Broad adult trauma measures** | | |
| Exposure to war/collective violence | 1.13 (0.87-1.45) | 1.15 (0.71-1.87) |
| Illness, accident and natural disaster | 1.06 (0.86-1.30) | 1.14 (0.76-1.71) |
| Household challenges | 0.60 (0.33-1.10) | 2.98 (1.47-6.05) |
| Abuse | 1.34 (0.90-2.02) | 0.36 (0.05-2.52) |

Supplementary Table 10. Hazard ratios and odds ratios for dementia incidence per additional adversity experienced using complete case data.

| **Sum adversity scores** | | | |
| --- | --- | --- | --- |
| **Total number of adversities** | | | |
| **Primary Analysis** | **HR** | **95% CI** | |
| Cox proportional hazards model | 1.06 | 1.00 | 1.13 |
| **Sensitivity Analysis** | **OR** | **95% CI** | |
| Logistic regression: no one censored developed dementia | 1.08 | 1.01 | 1.15 |
| Logistics regression: everyone censored developed dementia | 1.01 | 0.96 | 1.05 |
| **Number of adult adversities** | | | |
| **Primary Analysis** | **HR** | **95% CI** | |
| Cox proportional hazards model | 1.11 | 1.03 | 1.20 |
| **Sensitivity Analysis** | **OR** | **95% CI** | |
| Logistic regression: no one censored developed dementia | 1.13 | 1.03 | 1.23 |
| Logistics regression: everyone censored developed dementia | 1.03 | 0.97 | 1.10 |
| **Number of childhood adversities** | | | |
| **Primary Analysis** | **HR** | **95% CI** | |
| Cox proportional hazards model | 1.99 | 0.88 | 1.11 |
| **Sensitivity Analysis** | **OR** | **95% CI** | |
| Logistic regression: no one censored developed dementia | 1.02 | 0.90 | 1.16 |
| Logistics regression: everyone censored developed dementia | 0.97 | 0.90 | 1.05 |

Possible ranges of sum adversity scores are as follows: Total (0-25), Childhood (0-12), Adulthood (0-13). Primary analysis - Cox proportional hazards model of dementia incidence in association with adversity measures, adjusting for sex and ethnicity, using age as the time axis (HR). Sensitivity analysis: No one censored developed dementia – logistic regression of dementia incidence in association with adversity measures, adjusting for age, quadratic age and ethnicity assuming everybody lost to follow up did not develop dementia (OR). Sensitivity analysis: Everyone censored developed dementia – logistic regression of dementia incidence in association with adversity measures, adjusting for age, quadratic age and ethnicity assuming everybody lost to follow up did develop dementia (OR).

Supplementary Table 11. The association between broad adversity measures and dementia incidence using complete case data. Cox models were adjusted for sex, ethnicity and childhood SES. Age was used as the time axis.

| **Broad Measures** | | | |
| --- | --- | --- | --- |
| **Cox regression** | **HR** | **95% CI** | |
| **Adult adversities** | | | |
| Exposure to war/collective violence | 1.02 | 0.69 | 1.45 |
| Illness, accident and natural disaster | 0.86 | 0.68 | 1.09 |
| Household challenges | 0.96 | 0.60 | 1.52 |
| Economic hardship | 1.96 | 0.71 | 1.30 |
| Abuse | 1.78 | 1.21 | 2.63 |
| **Childhood adversities** |  |  |  |
| Exposure to war/collective violence | 1.28 | 0.98 | 1.67 |
| Illness, accident and natural disaster | 1.06 | 0.86 | 1.32 |
| Household challenges | 1.05 | 0.63 | 1.73 |
| Economic hardship | 1.27 | 0.98 | 1.64 |
| Abuse | 1.28 | 1.83 | 1.97 |

Supplementary Figure 1. Tetrachoric correlation matrix of individual adversity measures.

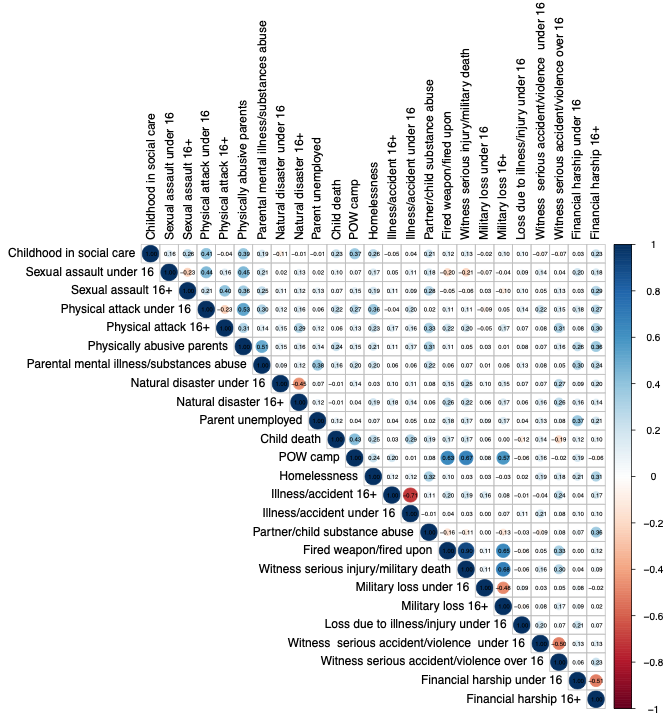

Supplementary Figure 2. Tetrachoric correlation matrix of broad adversity measures.

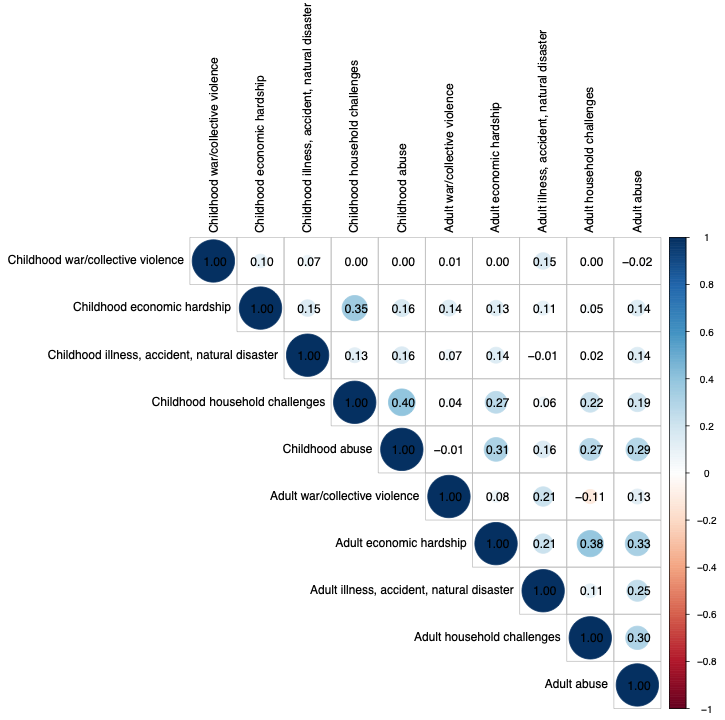

Supplementary Figure 3. Modelling the impact of increasing sum adversity score category, compared to having experienced no adverse experiences, on hazard/odds of dementia incidence.

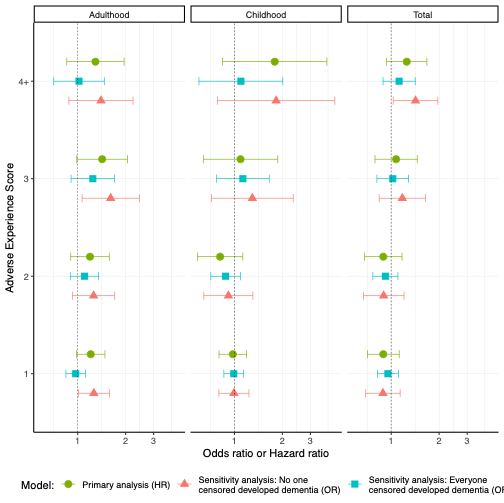

Primary analysis - Cox proportional hazards model of dementia incidence in association with adversity measures, adjusting for sex and ethnicity, using age as the time axis (HR). Sensitivity analysis: No one censored developed dementia – logistic regression of dementia incidence in association with adversity measures, adjusting for age, quadratic age and ethnicity assuming everybody lost to follow up did not develop dementia (OR). Sensitivity analysis: Everyone censored developed dementia – logistic regression of dementia incidence in association with adversity measures, adjusting for age, quadratic age and ethnicity assuming everybody lost to follow up did develop dementia (OR).
